# Students as Community Vaccinators: Implementation of a Service-Learning COVID-19 Vaccination Program

**DOI:** 10.1101/2022.01.14.22269312

**Authors:** Andrew R. Griswold, Julia Klein, Neville Dusaj, Jeff Zhu, Allegra Keeler, Erika L. Abramson, Dana Gurvitch

## Abstract

**Background:** Service-learning is an integral component of medical education. While the COVID-19 pandemic has caused massive educational disruptions, it has also catalyzed innovation in service-learning as real-time responses to pandemic-related problems. For example, the limited number of qualified providers was a potential barrier to local and national SARS-CoV-2 vaccination efforts. Foreseeing this hurdle, New York State temporarily allowed healthcare professional trainees to vaccinate, enabling medical students to support an overwhelmed healthcare system and contribute to the community. Yet, it was the responsibility of medical schools to interpret these rules and implement the vaccination programs. Here the authors describe a service-learning vaccination program directed towards underserved communities.

**Methods:** Weill Cornell Medicine (WCM) rapidly developed a faculty-led curriculum to prepare students to communicate with patients about the COVID-19 vaccines and to administer intramuscular injections. Qualified students were deployed to public vaccination clinics located in underserved neighborhoods across New York City in collaboration with an established community partner. The educational value of the program was evaluated with retrospective survey.

**Results:** Throughout the program, which lasted from February to June 2021, 128 WCM students worked at 103 local events, helping to administer 26,889 vaccine doses. Analysis of student evaluations revealed this program taught fundamental clinical skills, increasing comfort giving intramuscular injection from 2% to 100% and increasing comfort talking to patients about the COVID-19 vaccine from 30% to 100%. Qualitatively participants described the program as a transformative service-learning experience.

**Conclusion:** As new virus variants emerge, nations battle recurrent waves of infection, and vaccine eligibility expands to include children and boosters, the need for effective vaccination plans continues to grow. The program described here offers a novel framework that academic medical centers could adapt to increase vaccine access in their local community and provide students with a uniquely meaningful educational experience.

## INTRODUCTION

Service learning is an experiential educational approach that combines community engagement with academic coursework and personal reflection^1,2^. It is a core component of medical training and required for institutional accreditation by the Liaison Committee on Medical Education (LCME)^3,4^. Over the past two-years, the COVID-19 pandemic has disrupted the delivery of medical education and forced the role of medical students to evolve. Nevertheless, medical students have found ways to contribute to the overwhelmed health care system and serve their community in these uncertain times. While many initiatives occurred in non-patient facing roles^5-9^, some service-learning experiences incorporated direct patient contact. Most notably, perhaps, was the accelerated graduation and deployment of senior medical students as new physicians^10,11^. Students have also screened patients for COVID-19^12^, provided telemedicine support^13^, worked at patient call centers^14^, and facilitated the reopening of student run free-clinics^15^. Collectively these examples show that although medical students are generally accepted to be non-essential workers^16^; they do represent a large pool of untapped providers with tremendous potential.

The shortage of qualified healthcare professionals capable of administering vaccinations at the scale required early in the pandemic presented a unique service-learning opportunity to medical students with historic precedent. During the great influenza pandemic of 1918, American medical students assisted with the mass inoculation program^17^. Indeed, on January 8, 2021 New York expanded executive order 202.9 which enabled professional students to become eligible vaccinators given they met the established standards. Despite the guidance provided by this executive action, the development and execution of student programs remained the responsibility of academic institutions. We were particularly interested in how to create a sustainable program that serviced underrepresented communities while being a value-added experience for medical student volunteers. However, there was a dearth of scholarship about student-run vaccination clinics and no literature about a vaccination program of the scale and duration envisioned for COVID-19. Here we describe the design, implementation, and evaluation of our institution’s first of its kind student vaccinator program.

## MATERIALS AND METHODS

### Program Development

Following the executive order enabling students to serve as vaccinators, Weill Cornell Medicine began training students as volunteer vaccinators to staff community vaccination sites organized by the Community Healthcare Network (CHN), a federally qualified health center. Guidelines for student training were set by the NYS Department of Health (DOH) to include completion of at least one year of clinical training, vaccine specific online training modules, an in-person training session, and BCLS certification. Students who had completed the full foundational curriculum in addition to at least one clerkship were eligible for training. Student volunteers were required to be fully vaccinated or willing to take both vaccine doses at associated vaccine events. Students who did not meet the clinical curricular requirements and/or were not vaccinated were excluded from training and volunteering.

A skills checklist for the in-person skills demonstration was provided by the NYC.GOV website (https://www.immunize.org/catg.d/p7010.pdf). In-person skills training session were held in an Objective Structured Clinical Examination (OSCE) format familiar to medical students under supervision of an anesthesiology resident and/or attending physician. All NYS guidelines for Personal Protective Equipment (PPE) were strictly adhered to and measures were discussed regarding managing adverse outcomes. During the OSCE-style sessions, students were supervised conducting a complete mock COVID-19 vaccination encounter including patient education, vaccine preparation, and immunization administration of a saline intramuscular (IM) injection on a fellow classmate. Upon successful completion of all training components, students were added to the certified roster and became eligible to volunteer at vaccination events.

### Vaccination Events

Vaccination events occurred from February to July 2021. Patient eligibility was in accordance with NYS guidelines (**Figure 1A**). Each event had medical oversight by either an attending physician or nurse practitioner. Students were responsible for reconstituting vaccine doses, administering vaccinations, counseling patients regarding vaccine expectations, and appropriately documenting vaccinations. All these tasks were performed jointly by medical students, nurses from our partner clinic, and attending physicians and nurse practitioners. Data on vaccine recipients between were collected by Community Health Network. Demographic breakdowns of patients were calculated for Age, Sex at Birth, and Race/Ethnicity.

**Figure 1.**
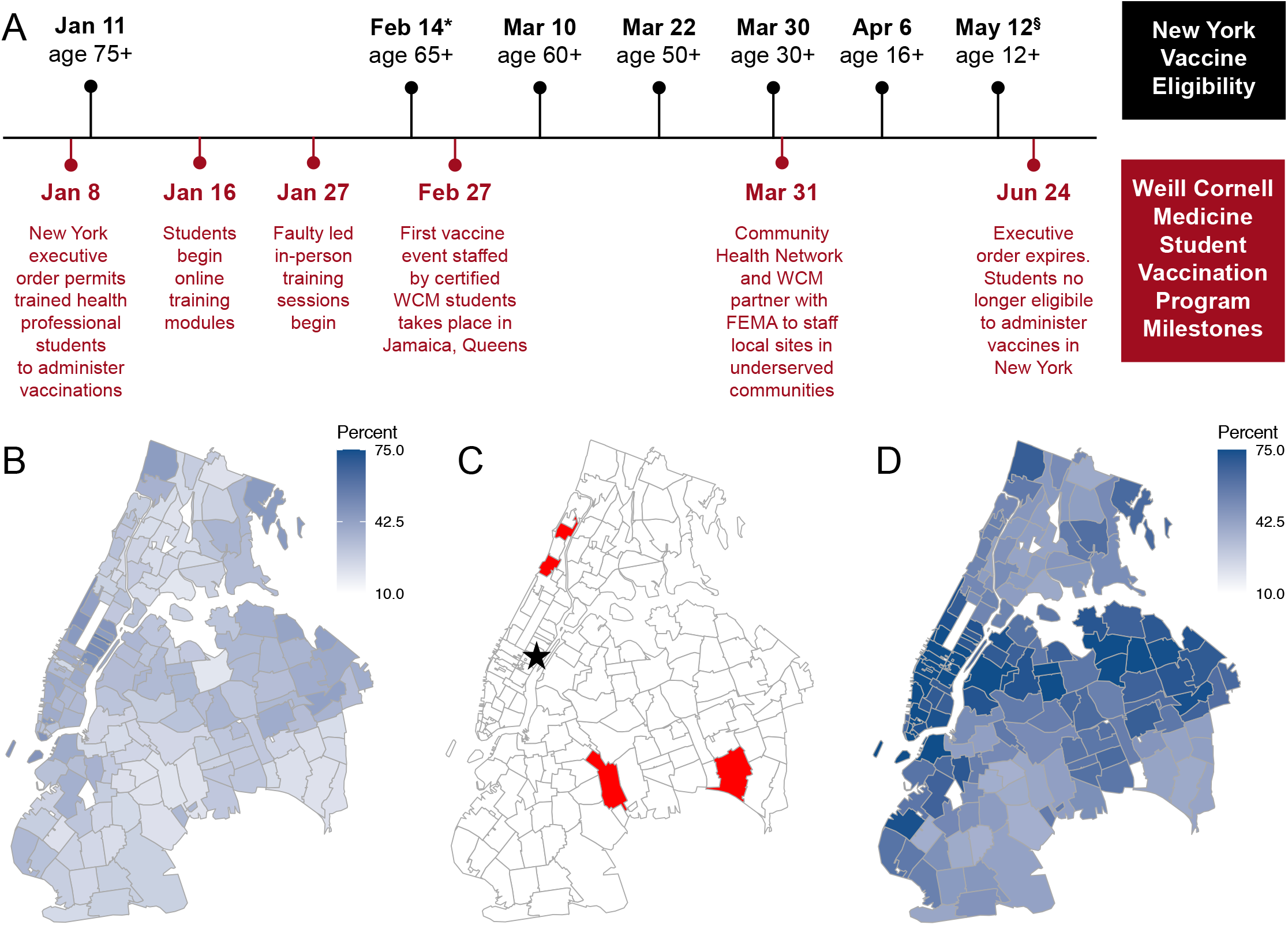
Timeline and map of student vaccination program in 2021 (A) New York State COVID-19 vaccine eligibility dates (top, black) superimposed with summary of key events of WCM vaccination program (bottom, red). Note timeline not drawn to scale. *Vaccine eligibility also extended to people with underlying medical conditions on February 14. ^§^People ages 12-15 were exclusively eligible for the Pfizer-BioNTech COVID-19 vaccine. (B) Heatmap of NYC vaccination status, at least one dose by March 26, 2021. (C) Locations of WCM vaccination program sites (red) and Weill Cornell Medicine (star). (D) Heatmap of NYC vaccination status at least one dose, by July 1, 2021, as colored in B.

Vaccination events took place throughout NYC at transient (pop-up) and stable (Federal Emergency Management Agency (FEMA)-funded) sites. The permanent locations were intentionally established in under-served and under-represented communities: Church of God in Brooklyn, Fort Washington Collegiate Church in Washington Heights, New Jerusalem Worship Center in Jamaica Queens and Convent Avenue Baptist Church in Harlem (**Figure 1B-D**). These sites were open on weeknights and weekends to minimizes barriers to access for working individuals. Transportation was provided for student volunteers. Community partners provided critical support including check-in and scheduling to translation assistance and custodial work. FEMA and NY Department of Health (NYDOH) delivered vital backing – ensuring an uninterrupted supply of vaccines, creating a digital infrastructure for tracking patient data, and providing the funds for much of this program.

### Retrospective Survey Development and Administration

The program was evaluated with a retrospective survey. Questions for medical student vaccinators were developed to address the following broad categories: quantify degree of involvement in the vaccination program, safety, skills development (hands-on and communication with patients), areas for improvement, and personal reflections. The survey was administered via RedCap and included quantitative questions about training and open-ended questions on value of experience. A copy of the survey can be found in **Supplemental Figure 1**. Students who participated in at least one event were provided with anonymous online survey link via their WCM email address (or personal email addresses for M4 students, all of whom graduated prior to distribution). Completion of the survey was optional.

### Ethical approval

Weill Cornell Medicine Human Research Compliance conducted a review of our protocol (#21-08023826) on September 23^rd^, 2021 and determined that the activities described did not constitute human subjects research. Therefore, IRB approval was not required for this study.

## RESULTS

Our program was unique in its ability to quickly train vaccinators, place students at vaccination sites and collect data regarding this service-learning opportunity, which became a unique curricular experience for WCM students that may serve as a model for similar service-learning efforts. The ever-changing landscape of State and Federal guidelines temporally guided many program milestones (**Figure 1A**). For example, within one month of the January executive order, WCM students had completed online modules and attended faculty-directed training sessions as described above. The first student-staffed vaccination event on February 27, occurred during a surge of vaccine demand corresponding to expansion of vaccine eligibility for adults 65 years of age and older and people with underlying medical conditions. At this first event, students helped inoculate 176 patients with no major adverse events, providing proof of principle that the student vaccination program was safe and effective.

### Student Participants

Based on the inclusion criteria, 324 WCM students were eligible for vaccine training. Of these, 128 completed training and became certified vaccinators (40%). These students were from both MD and MD-PhD programs and tended to be in the later years of training (**Table 1**). 80 students (63% of certified) participated in at least one or more vaccination event.

**Table 1.** Demographics of students certified as vaccinators and patients vaccinated through the Program.

| **Student Vaccinator Characteristic** | **% (N = 128)** |
| --- | --- |
| *Program* |  |
| MD | 68 (87) |
| MD-PhD | 32 (41) |
| *Class Year* |  |
| 2 | 13 (16) |
| 3 | 34 (44) |
| 4+ | 53 (68) |
| **Patient Demographic** | **% (N = 14,293)** |
| *Age* |  |
| <18 | 17 (2451) |
| 18-24 | 12 (1683) |
| 25-34 | 18 (2548) |
| 35-44 | 18 (2555) |
| 45-54 | 15 (2147) |
| 55-64 | 13 (1796) |
| 65+ | 7 (1057) |
| *Sex at Birth* |  |
| Female | 50 (7145) |
| Male | 48 (6822) |
| *Race/Ethnicity* |  |
| White | 10 (1390) |
| Black | 30 (4345) |
| Asian | 10 (1365) |
| Hispanic | 39 (5557) |
| Other | 9 (1287) |

### Patient Demographics

Students helped to inoculate 14,293 individuals with 26,889 vaccine doses (**Table 1**). Patients were relatively equally distributed in sex at birth, with slightly more females (7145) than males (6822). The majority of the patients (9237) were younger than 45 in age. While this is contrary to national trends that elder individuals are more likely to be vaccinated^18^, it likely stems from the timing of when recurring FEMA sites were established (March 31). For example, individuals ages 30-49 only become eligible for vaccines on March 30, whereas older people had been eligible for vaccines for up to 3 months. Our program was successful in reaching racial and ethnic minorities with 30% of patients identifying as Black and 39% of patients identifying as Hispanic. Notably, only 10% of the patients reached by this program identified as White.

### Evaluation of the Vaccination Program

The effect of the program as a service-learning opportunity for student participants was evaluated through a retrospective survey. Fifty students (50/80, 63% response rate) completed the optional post-volunteer survey. A summary of the results as well as representative excerpts are shown in **Table 2** and **Table 3**. Overall, the training was well received, with all respondents noting that they felt adequately prepared to participate. Training was able to increase the percent of students feeling comfortable administering IM injections from 2% to 76%. The experience of attending vaccination events, however, was essential in solidifying clinical skills. Notably, 100% of students indicated they were comfortable talking to patients about the COVID-19 vaccine and administering IM injections after the vaccination program. Similarly, participation in this program increased student comfort talking to patients about the COVID-19 vaccine. Together these data show that this service-learning experience provided educational value and boosted clinical confidence.

**Table 2.** Summary of student evaluations for vaccination experience.

| **Program Evaluation Question** | **% (N = 50)** |
| --- | --- |
| Did you feel you had adequate training and preparation to participate? |  |
| Yes | 100% (49) |
| No | 0% (0) |
| Did you feel protected wearing appropriate PPE and adhering to NYS guidelines? |  |
| Yes | 100% (50) |
| No | 0% (0) |
| How comfortable did you feel with IM injections prior to training? |  |
| Not at all or somewhat comfortable | 98% (49) |
| Comfortable or very comfortable | 2% (1) |
| How comfortable did you feel with IM injections post-training? |  |
| Not at all or somewhat comfortable | 24% (12) |
| Comfortable or very comfortable | 76% (38) |
| How comfortable do you feel with IM injections now [i.e. post vaccination program)? |  |
| Not at all or somewhat comfortable | 0% (0) |
| Comfortable or very comfortable | 100% (49) |
| How comfortable did you feel talking to patients about the COVID-19 vaccine prior to volunteering? |  |
| Not at all or somewhat comfortable | 70% (35) |
| Comfortable or very comfortable | 30% (15) |
| How comfortable do you feel talking to patients about the COVID-19 vaccine after volunteering? |  |
| Not at all or somewhat comfortable | 0% (0) |
| Comfortable or very comfortable | 100% (50) |
| Would you sign up for similar volunteer efforts in the future? |  |
| Yes | 100% (50) |
| No | 0% (0) |

**Table 3.**
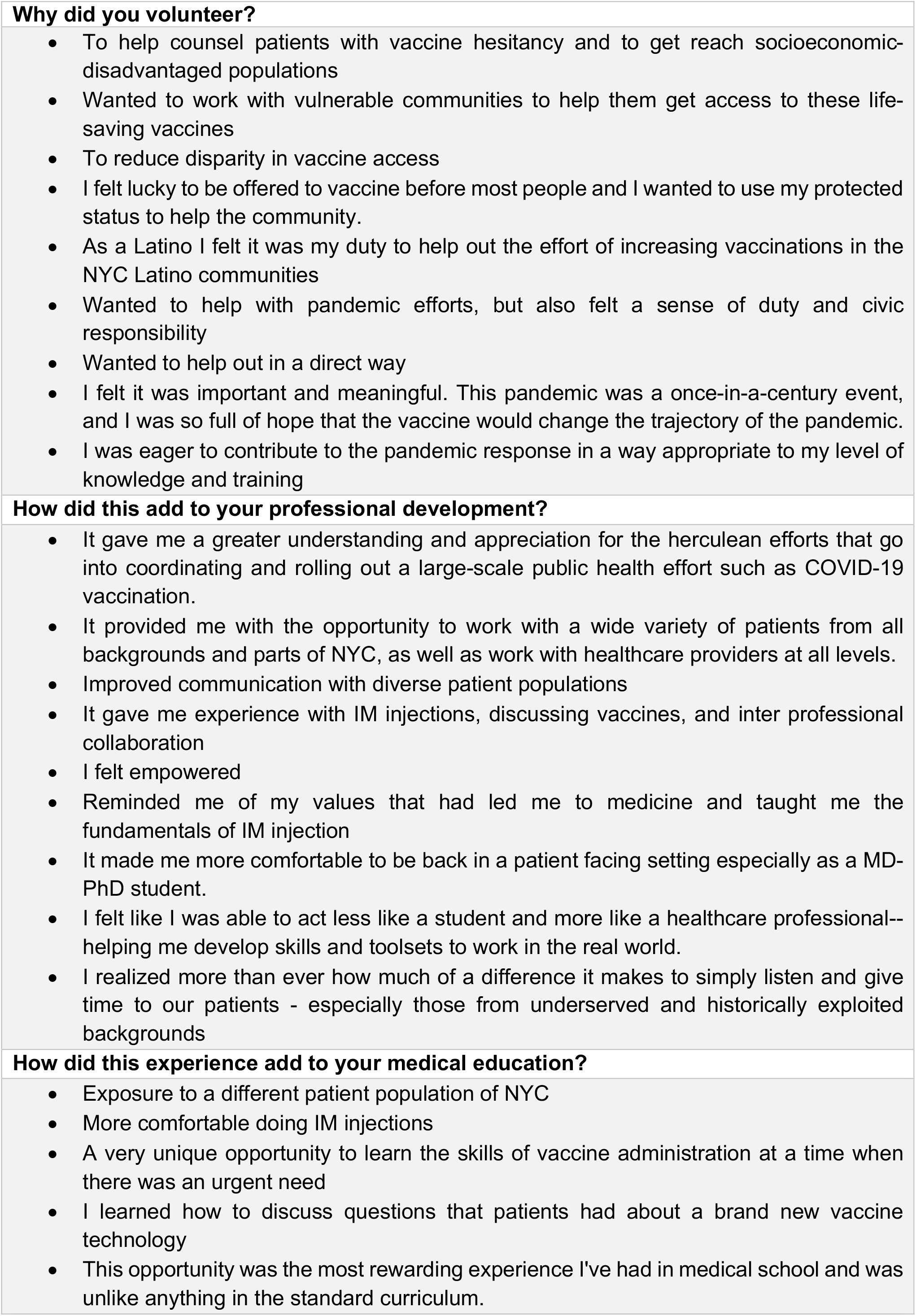
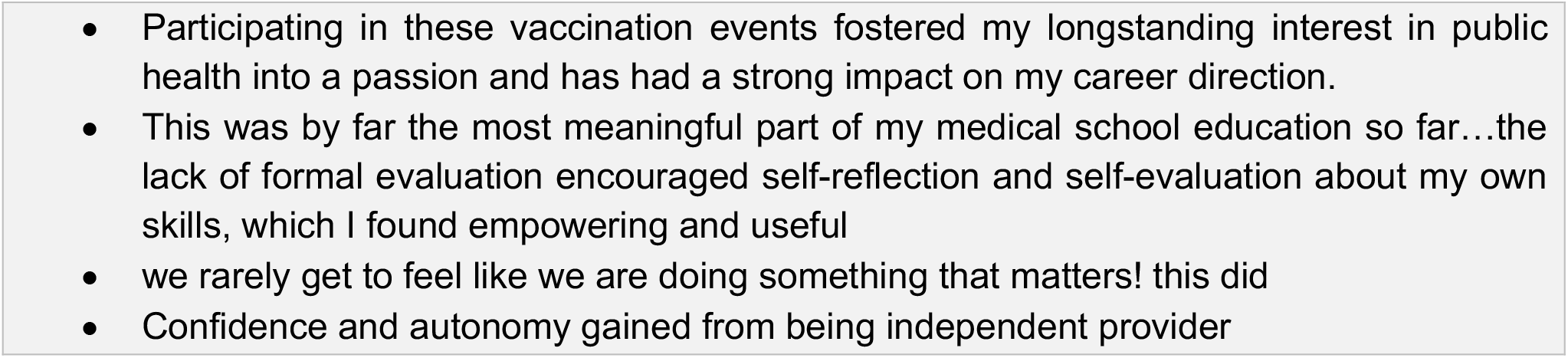
Student perceptions of the vaccination program

Inspection of qualitative responses further indicate that the vaccination program was an opportunity for professional growth. When asked why they volunteered, students indicated desires to work with vulnerable communities, reduce disparity in vaccine access, feeling a sense of civic responsibility to help with pandemic efforts and wanting to help pandemic efforts in a direct way. When describing how volunteering as a vaccinator contributed to their professional development, student responses ranged from improving communication with diverse patient populations to better understanding how large-scale public health efforts are coordinated an executed. Finally, when asked how this experience contributed to their medical education students gave many reasons, including fostering an interest in public health and gaining confidence and experience functioning as an independent provider. Multiple students indicated that this was the most meaningful part of their medical education to date.

## DISCUSSION

Here we discuss the development, implementation, and evaluation of an innovative service-learning COVID-19 vaccination program. While there is growing body of literature about medical student contributions to the COIVD-19 response^6-10,12,15,19-28^, this is the first paper to describe an initiative that trained students as vaccinators during an actively developing global pandemic.

The program met its goals, to expand access to the COVID-19 vaccine and to provide a valuable service-learning experience for medical students. Crucial to this success were the three key-stakeholder groups – medical students, community partners and the government – each of whom made unique contributions to the program. Medical students formed the backbone of the program workforce by increasing the number of eligible vaccinators. They were trained providers, eager to volunteer, and altruistic with their motives. Collaborating with established community partner meant efforts to be directed towards delivering care to vulnerable communities in need of vaccination infrastructure without extensive relationship building. Finally, government bodies provided vital support by guaranteeing a reliable supply of vaccines, passing policies which enabled students to become qualified vaccinators and providing key financial support required to sustain a long-term program. While the collaborative approach was overwhelming positive, one limitation was that a third-party managed the digital infrastructure tracking vaccines. As such, we were unable to retrospectively analyze vaccine data based on the provider (i.e. medical student, nurse, physician associate, or physician). Nevertheless, we believe that this three-part model integrating students, community partners and government agencies, can be utilized to enhance community-outreach programs at our institution and can be emulated for future service-based learning programs at medical schools across the country.

Given the emergence of new SARS-CoV-2 variants (B.1.1.529 – Omicron, B.1.617.2 – Delta) and evolving nature of the COVID-19 pandemic the need for COVID-19 vaccinations remains high. Primary doses are available to children younger than 18, booster shots are available to most adults, and at least 20% of the population eligible to receive the vaccine has yet to receive a single dose. As the Public Readiness and Emergency Preparedness (PREP) Act continues to allow medical students to serve as vaccinators, institutions around the country have the potential to help fulfill these needs. Here, we provide a skills-based, checklist-guided approach to ensure competency amongst medical students that can be easily adapted and implemented by all medical schools. Competency can be readily identified by most supervising attendings or house staff from the affiliated academic medical center. Once trained, students can help administer vaccine doses at both their home medical institutions’ clinical locations, as well as at partner clinic locations, where they would remain under the same level of supervision as they would when completing training clerkships or sub-internships. We hope this meaningful and impactful service-learning model will be readily adopted.

## Data Availability

All data produced in the present study are available upon reasonable request to the authors.

## ACKNOWLEDGEMENTS

The authors wish to thank The WCM Student Vaccination Consortium (Abderhman Abuhashem, Evan Balmuth, Nicolas Blobel, Jeremy Chang, Hannah Chen, Lala Tanmoy Das, Deeksha Deep, Gregory Han, Margaret Herre, Iryna Ivasyk, Seong Jang, Jerry Lee, Katie Liu, Allison Moyer, Natalie Nguyen, Anthony Palillo, Nicola Pereira, Harlan Pietz, Daniel Poston, Devin Ray, Rachel Rosengard, Daniel Shinn, Amber Simmons, Aleksandr Talishinsky, Jacqueline Tran, Stefan Torborg, Marcos Lu Wang, Eric Zheng and Andrew Zhu), Yoon Kang, Kelli Ruttle, Shelby Badani, Harmandeep Singh, Robert White, Jay Swathirajan, students who volunteered at vaccination events, the medical oversight providers, Community Healthcare Network, and The Clinical & Translational Science Center.

## FUNDING/SUPPORT

This work was supported by the National Institutes of Health (F30 CA243444 to A.R.G., F30 HL156496 to N.D., T32 GM007739-Andersen to A.R.G and N.D., and UL1TR002384 to Clinical & Translational Science Center). The content is solely the responsibility of the authors and does not necessarily represent the official views of the National Institutes of Health.

## DISCLOSURE STATEMENT

The authors report there are no competing interests to declare.

## SUPPLEMENTAL MATERIALS

Supplemental data for this article can be accessed online.

## DATA AVAIBILITY

Not applicable

---

**Supplemental Figure 1.**
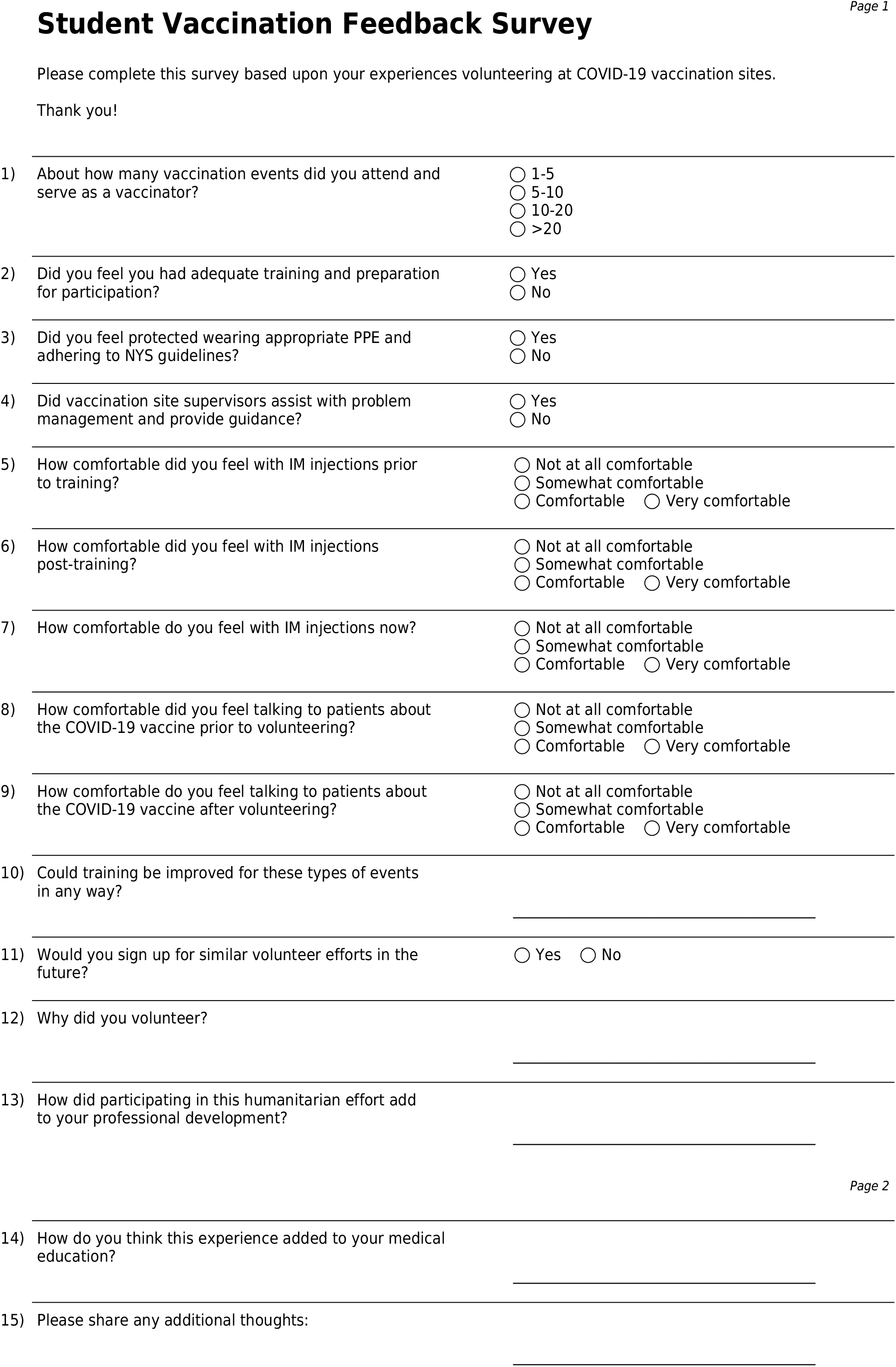
Student Vaccination Feedback Survey

## Notes

### Competing Interest Statement

The authors have declared no competing interest.

### Author Declarations

Weill Cornell Medicine Human Research Compliance conducted a review of our protocol (#21-08023826) on September 23rd, 2021 and determined that the activities described did not constitute human subjects research. Therefore, IRB approval was not required for this study.

## REFERENCES

1. Stewart T, Wubbena ZC. A systematic review of service-learning in medical education: 1998-2012. Teach Learn Med. 2015;27(2):115–22. doi:10.1080/10401334.2015.1011647

2. Hunt JB, Bonham C, Jones L. Understanding the goals of service learning and community-based medical education: a systematic review. Acad Med. Feb 2011;86(2):246–51. doi:10.1097/ACM.0b013e3182046481

3. LCME. Functions and Structure of a Medical School: Standards for Accreditation of Medical Education Programs Leading to the MD Degree. 2021.

4. Nguemeni Tiako MJ, Johnson SF, Nkinsi NT, Landry A. Normalizing Service Learning in Medical Education to Sustain Medical Student-Led Initiatives. Acad Med. Sep 28 2021;doi:10.1097/ACM.0000000000004432

5. Zhou B, Calkins C, Jayaraman T, et al. Implementing Value-Added Medical Education. Academic Medicine. 2021; Publish Ah doi:10.1097/acm.0000000000004160

6. Kochis M, Goessling W. Learning during and from a Crisis: The Student-Led Development of a COVID-19 Curriculum. Academic Medicine. 2021:399–401. doi:10.1097/ACM.0000000000003755

7. Alshak MN, Li HA, Wehmeyer GT. Medical Students as Essential Frontline Researchers during the COVID-19 Pandemic. Lippincott Williams and Wilkins; 2021. p. 964–966.

8. Villela EFdM, de Oliveira FM, Leite ST, Bollela VR. Student engagement in a public health initiative in response to COVID-19. Medical Education. 2020;54(8):763–764. doi:10.1111/MEDU.14199

9. Long N, Wolpaw DR, Boothe D, et al. Contributions of Health Professions Students to Health System Needs during the COVID-19 Pandemic: Potential Strategies and Process for U.S. Medical Schools. Academic Medicine. 2020:1679–1686. doi:10.1097/ACM.0000000000003611

10. Flotte TR, Larkin AC, Fischer MA, et al. Accelerated Graduation and the Deployment of New Physicians during the COVID-19 Pandemic. Lippincott Williams and Wilkins; 2020. p. 1492–1494.

11. “Itching to get back in”: Medical students graduate early to join the fight. https://www.aamc.org/news-insights/itching-get-back-medical-students-graduate-early-join-fight.

12. Nanette P, Ribault S, Peyrot S, et al. Medical student engagement in a massive COVID-19-screening programme. Medical Education. 2021;55(11):1299–1300. doi:10.1111/MEDU.14620

13. Soled D, Goel S, Barry D, et al. Medical Student Mobilization during a Crisis: Lessons from a COVID-19 Medical Student Response Team. Academic Medicine. 2020:1384–1387. doi:10.1097/ACM.0000000000003401

14. Tsang VWL, Yu A, Haines MJ, et al. Transforming Disruption into Innovation. Academic Medicine. 2021; Publish Ah doi:10.1097/acm.0000000000004156

15. Weiss C, Traczuk A, Motley R. Reopening a Student-Run Free Clinic During the COVID-19 Pandemic to Provide Care for People Experiencing Homelessness. Acad Med. Oct 19 2021;doi:10.1097/ACM.0000000000004480

16. Menon A, Klein EJ, Kollars K, Kleinhenz ALW. Medical students are not essential workers: Examining institutional responsibility during the COVID-19 pandemic. Academic Medicine. 2020;95(8):1149–1151. doi:10.1097/ACM.0000000000003478

17. Crosby AW. America’s forgotten pandemic: The influenza of 1918, second edition. Cambridge University Press; 2003:1–337.

18. U.S. COVID-19 vaccine tracker. Mayo Clinic. 2021. https://www.mayoclinic.org/coronavirus-covid-19/vaccine-tracker

19. Thomas AT, Gilja S, Sikka N, et al. Peer-to-peer COVID-19 medical curriculum development during the pandemic. Medical Education. 2021;55(11):1302–1303. doi:10.1111/MEDU.14639

20. Klasen JM, Meienberg A, Bogie BJM. Medical student engagement during COVID-19: Lessons learned and areas for improvement. Medical Education. 2021;55(1):115–118. doi:10.1111/MEDU.14405

21. Matthew D, Eftychiou L, French C, Hare A. ‘Just in time’ rapid learning during COVID-19. Medical Education. 2021;55(11):1300–1301. doi:10.1111/MEDU.14641

22. Castro MRH, Calthorpe LM, Fogh SE, et al. Lessons from learners: Adapting Medical Student Education During and Post-COVID-19. Academic Medicine. 2021;doi:10.1097/ACM.0000000000004148

23. Gordon M, Kim HW, Kennedy W, Bartolo IMD. Utilising medical students as wellness coaches during the CoVID-19 pandemic. Medical Education. 2021;55(11):1306–1307. doi:10.1111/MEDU.14617

24. Shibu A. Medical student engagement during the COVID-19 pandemic—A student perspective. Medical Education. 2021;55(6):768–768. doi:10.1111/MEDU.14474

25. Grilo SA, Catallozzi M, Desai U, et al. Columbia COVID-19 Student Service Corps: Harnessing student skills and galvanizing the power of service learning. FASEB BioAdvances. 2021;3(3):166–174. doi:10.1096/FBA.2020-00105

26. Magklara E, Angelis S, Solia E, et al. The Role of Medical Students During COVID-19 Era. A Review. Acta Bio Medica : Atenei Parmensis. 2021;92(1):2021032–2021032. doi:10.23750/ABM.V92I1.10873

27. Schuiteman S, Ibrahim NI, Hammoud A, Kruger L, Mangrulkar RS, Daniel M. The Role of Medical Student Government in Responding to COVID-19. Academic Medicine. 2021:62–67. doi:10.1097/ACM.0000000000003542

28. Edelman DS, Desai UA, Soo-Hoo S, Catallozzi M. Responding to hospital system and student curricular needs: COVID-19 Student Service Corps. Medical Education. 2020;54(9):853–854. doi:10.1111/MEDU.14243

